# Developing an ICU Mortality Risk Prediction Model for Acute Myocardial Infarction Patients Based on Machine Learning

**DOI:** 10.1101/2025.08.31.25334808

**Authors:** Ying-mei Xiao, Juan Zhang, Mao-juan Wang

## Abstract

**Purpose:** Patients with acute myocardial infarction (AMI) are in a critical condition, facing a high risk of death in the intensive care unit (ICU) with significant individual differences. The aim of this study is to integrate clinical data using machine learning algorithms to construct a model for predicting the risk of death in ICU for AMI patients, thereby providing clinicians with an objective risk assessment tool.

**Patients and methods:** This study is a retrospective study that included 2285 patients from the Medical Information Mart for Intensive Care (MIMIC)-IV database. The primary outcome was in-hospital mortality of ICU patients with acute myocardial infarction. Univariate analysis was performed to screen statistically significant variables, and Lasso regression was used to further identify independent influencing factors that were significantly related to the risk of death. Based on the screened variables, we developed multiple machine learning(ML) models and evaluated their predictive efficiency for the risk of death in ICU patients with acute myocardial infarction.

**Results:** A total of 2285 ICU-admitted AMI patients were included, and 613 AMI patients died in the ICU. After the screening process, a total of 15 clinical characteristic variables were included in developing logistic regression models and ten ML models (each model utilized the same 15 clinical characteristics), and the table outlines the predictive performance of these models. According to the area under the Area Under the Curve(AUC), the 3 prediction models showed good predictive efficacy for AMI ICU mortality risk. The CatBoostTEST model AUC = 0.78, the LGBMTEST model AUC = 0.766, and the RFTEST model AUC = 0.755.

**Conclusion:** Machine learning models provide an excellent tool for predicting the risk of death from myocardial infarction in the ICU, laying the groundwork for potential improvements in clinical decision-making and patient outcomes. Machine learning models offer a good predictive tool for the mortality risk of AMI in the ICU, enabling early and accurate identification of high-risk patients, timely implementation of targeted interventions, and ultimately reducing the ICU mortality rate of AMI patients.

## Introduction

AMI is a cardiovascular emergency characterized by rapid onset, swift progression, and high mortality rates, posing a significant threat to global public health[1–2]. According to the World Health Organization, over 17 million individuals succumb to cardiovascular diseases annually worldwide, representing 31% of all deaths, with AMI being a leading contributor[3]. In our country, the incidence of AMI is escalating annually due to an aging population, changing lifestyles, and heightened social stress[4]. Additionally,the age of onset is progressively decreasing, resulting in a 5% increase in heart attacks among young people, thereby placing substantial strain on individual health and the social medical system[5]. ICUs frequently serve as primary treatment and death sites for critically ill AMI patients. Research suggests that the mortality rate of AMI patients in ICUs significantly surpasses that in general wards, making it a major contributor to in-hospital deaths[6–7]. Consequently, timely and accurate identification of AMI patients in ICUs who are at high risk of death, and the implementation of targeted intervention measures, are crucial for enhancing patient outcomes and reducing mortality rates in ICUs.

Presently, widely-utilized myocardial infarction risk scoring systems in clinical practice, such as GRACE and TIMI scores, exhibit certain predictive value[8–9]. However, their predictive efficiency has significant limitations, particularly regarding mortality risk prediction in ICU settings[10–11]. These scores are primarily based on linear regression models that depend on a limited set of clinical variables, thus failing to fully encapsulate the complex physiological changes in patients. Furthermore, these traditional scoring systems lack the adaptability to dynamically changing clinical data within the ICU environment, hindering their ability to meet precision and personalized diagnosis and treatment needs. In recent years, machine learning technology has demonstrated immense potential in the field of medical predictive modeling[12–13]. This technology can efficiently process large-scale, multi-source heterogeneous clinical data. This data includes real-time monitored vital signs in the ICU, laboratory indicators, and diagnostic and therapeutic measures. The technology can automatically uncover deep associations between complex features and build models with superior predictive accuracy and robustness. Preliminary studies in the field of cardiovascular diseases have confirmed the superiority of machine learning in predicting adverse events such as heart failure[14–15] and arrhythmias[16].

This study endeavors to develop a predictive model for the risk of mortality among patients with AMI admitted to ICU, utilizing machine learning algorithms. By synthesizing multi-dimensional clinical data from patient admissions to the ICU—encompassing basic information, vital signs, laboratory test results, comorbidities, and treatment histories—the study employs sophisticated machine learning techniques to ascertain the optimal predictive model. The objective is to furnish ICU clinicians with a refined and efficient tool for risk stratification of ICU mortality, thereby facilitating the early detection of high-risk AMI patients within the ICU. This, in turn, aims to enhance clinical decision-making and resource allocation within the ICU, ultimately diminishing the mortality rate of AMI patients and augmenting patient survival outcomes.

## Methods

### Study design

We analyzed the data derived from the MIMIC-IV database ((version 3.0). This database was established by the Computational Physiology Lab at the Massachusetts Institute of Technology, the Beth Israel Deaconess Medical Center at Harvard Medical School, and the Philips Healthcare. Data from 2008 to 2022 were covered.

### Study population

Inclusion criteria: (1) patients aged ≥18 years old; (2) according to the fourth edition of the global myocardial infarction definition criteria, myocardial infarction is defined as[17]: elevated myocardial enzyme markers (cTn), accompanied by at least one of the following: clinical symptoms of myocardial ischemia; new myocardial ischemic changes on electrocardiogram; pathological Q waves on electrocardiogram; imaging evidence of new loss of myocardial viability or regional wall motion abnormalities. Exclusion criteria: (1) patient age <18 years old; (2) hospitalization time less than 24 h; (3) the data is missing up to 30%.

### Data extraction and processing

Data were sourced from the MIMIC-IV database (version 3.0), which included the following patient information with acute myocardial infarction: (1) demographics; (2) initial vital signs, namely noninvasive mean blood pressure, noninvasive diastolic blood pressure, heart rate, and blood oxygen saturation; (3) initial laboratory test results, including absolute lymphocyte count, hematocrit, hemoglobin, platelet count, red cell distribution width, red blood cells, white blood cells, absolute neutrophil count, lactate, creatine kinase isoenzymes, lactate dehydrogenase, troponin T, international normalized ratio, clotting time, albumin, anion gap, total calcium, blood glucose, blood sodium, blood potassium, alanine transaminase, aspartame acid transaminase, total bilirubin, direct bilirubin, indirect bilirubin, creatinine, urea nitrogen, and fluid balance; (4) disease severity scores, specifically Sequential Organ Failure Assessment(SOFA) score and Acute Physiology and Chronic Health Evaluation II(APACHEII) score; (5) comorbidities such as sepsis and acute kidney injury; and (6) therapeutic interventions, which include mechanical ventilation, blood purification, extracorporeal membrane oxygenation (ECMO), intraaortic balloon counterpulsation (IABP), and vasoactive drugs. The MIMIC-IV database often contains missing values. Conflicting values were apparent in abnormal and continuous variables, and these were treated as missing values if their absence exceeded 30% of the dataset. If the absence was less than 30%, fixed value imputation was used to fill these data gaps. All data extraction processes were executed using R software (version 4.4.1).

### Ethics approval

This study utilized the Medical Information Mart for Intensive Care (MIMIC-IV) database, a publicly available, de-identified database. The creation and publication of MIMIC-IV were approved by the Institutional Review Boards (IRB) of the Massachusetts Institute of Technology (MIT) and Beth Israel Deaconess Medical Center (BIDMC). As this study involved the analysis of pre-existing, anonymized data, separate ethics approval for this specific analysis was not required. All researchers completed the required CITI training and signed the Data Use Agreement to access the database.

### Statistical analysis

This study utilizes the intensive care unit database and employs machine learning techniques to develop predictive models. For quantitative data that follows a normal distribution, the mean is used as a descriptive measure. In cases where the data distribution is skewed, the interquartile range (IQR) serves as the descriptive statistic. Qualitative data are described using the composition ratio. Quantitative data are subjected to statistical significance tests such as the t-test or the Mann-Whitney rank-sum test. The chi-square test is employed when comparing two or more composition ratios of qualitative data.Variables showing statistically significant differences in univariate analysis are incorporated into the LASSO regression to identify those related to the risk of in-hospital death from acute myocardial infarction. The predictive power of these identified risk factors is then evaluated and confirmed using various machine learning models.

## Results

### Baseline characteristics

This study is based on the MIMIC-IV database, including a total of 2285 patients with acute myocardial infarction treated in the ICU. Among them, there were 613 cases of ICU in-hospital death, and 1672 cases of survival, with an ICU mortality rate of 26.83%. The patient screening process is detailed in Fig. 1. Based on whether the patient died in the ICU, they were divided into the death group (613 cases) and the survival group (1672 cases). The overall male patients were 805 (35.23%), and the female patients were 1480 (64.77%). The age of the enrolled patients was 70.32 ± 13.20 years.The baseline characteristics of patients with myocardial infarction are presented in Table 1.

**Figure 1.**
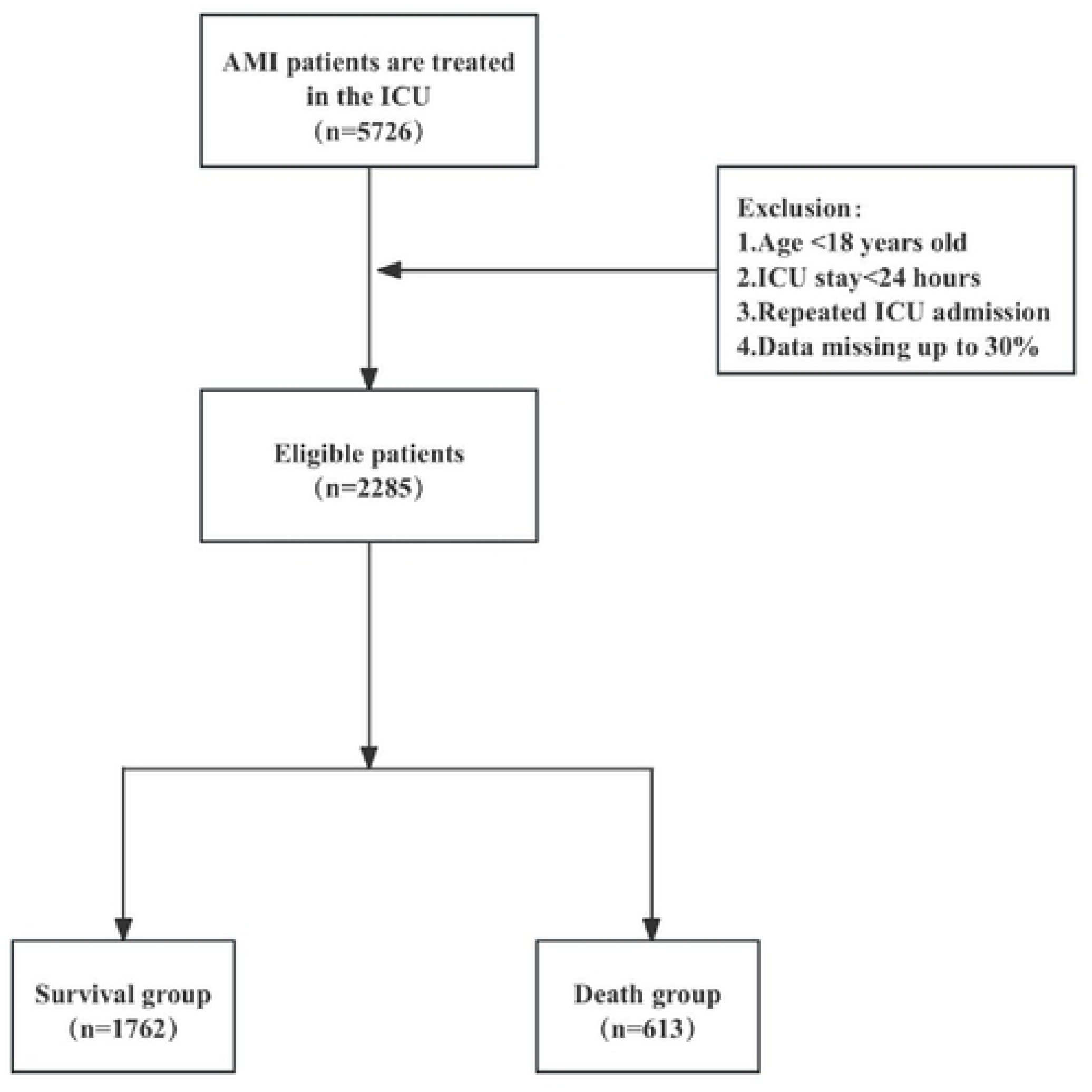
Screening flowchart of patients with AMI.

**Table 1.** Clinical characteristics of AMI admitted to ICU.

| Variable | Overall | Survival group<br>n= 1672 | Death group<br>n=613 | P |
| --- | --- | --- | --- | --- |
| Age, years (Mean±SD) | 70.32 ± 13.20 | 26.79±8.166 | 29.01±8.199 | <0.001 |
| Sex, N (%) |  |  |  |  |
| Males | 805.00 (35.23%) | 576.00 (34.45%) | 229.00 (37.36%) | 0.197 |
| Females | 1,480.00 (64.77%) | 1,096.00 (65.55%) | 384.00 (62.64%) |  |
| Initial vital signs(Mean±SD) |  |  |  |  |
| NMBP (mmHg) | 82.59 ± 18.96 | 83.49 ± 18.46 | 80.13 ± 20.08 | <0.001 |
| NDBP (mmHg) | 69.72 ± 18.89 | 70.47 ± 18.39 | 67.67 ± 20.06 | 0.003 |
| Heart rate (Beats per minute) | 90.24 ± 20.84 | 89.53 ± 20.94 | 92.17 ± 20.45 | 0.007 |
| Spo2 (%) | 96.17 ± 4.93 | 96.38 ± 4.72 | 95.59 ± 5.42 | 0.001 |
| Laboratory Examinations (Mean ± SD) |  |  |  |  |
| Absolute lymphocyte count (*10<sup>9</sup>/L) | 1.68 ± 9.35 | 1.80 ± 10.88 | 1.35 ± 1.70 | 0.096 |
| Hematocrit (mg/L) | 32.46 ± 7.37 | 32.65 ± 7.26 | 31.95 ± 7.62 | 0.049 |
| Platelet count (*10<sup>9</sup>/L) | 204.81 ± 105.44 | 208.85 ± 104.73 | 193.78 ± 106.64 | 0.003 |
| Hemoglobin (*10<sup>10</sup>/L) | 10.50 ± 2.51 | 10.60 ± 2.51 | 10.23 ± 2.51 | 0.002 |
| Rdw (%) | 15.38 ± 2.64 | 15.12 ± 2.51 | 16.08 ± 2.84 | <0.001 |
| Red blood cells count (*10<sup>9</sup>/L) | 3.54 ± 0.85 | 3.57 ± 0.85 | 3.45 ± 0.87 | 0.003 |
| White blood cells count (*10<sup>9</sup>/L) | 14.36 ± 9.98 | 14.06 ± 10.42 | 15.17 ± 8.59 | 0.010 |
| Absolute neutrophil count (*10<sup>9</sup>/L) | 11.73 ± 7.27 | 11.31 ± 7.14 | 12.90 ± 7.49 | <0.001 |
| Lactate (mmol/L) | 2.72 ± 2.50 | 2.32 ± 1.85 | 3.81 ± 3.51 | <0.001 |
| Creatine kinasemb isoenzyme (U/L) | 51.41 ± 96.95 | 48.00 ± 92.41 | 60.72 ± 107.92 | 0.010 |
| lactate dehydrogenase (IU/L) | 696.81 ± 1,208.93 | 580.68 ± 990.19 | 1,013.58 ± 1,624.80 | <0.001 |
| Troponin T (ug/L) | 1.77 ± 3.75 | 1.58 ± 3.28 | 2.28 ± 4.75 | <0.001 |
| INR | 1.60 ± 0.96 | 1.53 ± 0.88 | 1.79 ± 1.13 | <0.001 |
| Pt (s) | 17.44 ± 10.43 | 16.69 ± 9.61 | 19.51 ± 12.16 | <0.001 |
| Albumin (g/L) | 3.06 ± 0.60 | 3.13 ± 0.58 | 2.87 ± 0.62 | <0.001 |
| Alanine aminotransferasealt (U/L) | 175.78 ± 601.81 | 142.66 ± 526.10 | 266.10 ± 764.70 | <0.001 |
| Asparate aminotransferase (U/L) | 317.47 ± 1,121.98 | 250.39 ± 1,005.06 | 500.44 ± 1,376.28 | <0.001 |
| Total bilirubin(mg/dl) | 1.17 ± 2.58 | 0.96 ± 1.85 | 1.74 ± 3.90 | <0.001 |
| Anion gap(mmol/L) | 16.40 ± 5.13 | 15.75 ± 4.59 | 18.17 ± 6.05 | <0.001 |
| Total calcium(mmol/L) | 8.33 ± 0.86 | 8.35 ± 0.81 | 8.26 ± 1.00 | 0.051 |
| Glucose(mg/dl) | 177.72 ± 105.94 | 171.71 ± 103.94 | 194.12 ± 109.64 | <0.001 |
| Potassium (mmol/L) | 4.39 ± 0.83 | 4.36 ± 0.82 | 4.48 ± 0.87 | 0.003 |
| Sodium (mmol/L) | 137.96 ± 5.60 | 138.01 ± 5.48 | 137.82 ± 5.92 | 0.489 |
| Creatinine(mg/dl) | 2.10 ± 2.17 | 1.97 ± 2.17 | 2.47 ± 2.13 | <0.001 |
| Urea nitrogen(mg/dl) | 37.43 ± 27.97 | 34.92 ± 26.71 | 44.26 ± 30.12 | <0.001 |
| Balance (ml) | 2,026.37 ± 19,217.42 | -135.54 ± 18,534.04 | 7,923.14 ± 19,818.04 | <0.001 |
| SOFA | 6.85 ± 3.92 | 6.08 ± 3.58 | 8.96 ± 4.04 | <0.001 |
| APACHE II | 21.68 ± 8.11 | 20.06 ± 7.48 | 26.09 ± 8.11 | <0.001 |
| IABP, N (%) |  |  |  |  |
| No | 2,003.00 (87.66%) | 1,464.00 (87.56%) | 539.00 (87.93%) | 0.812 |
| Yes | 282.00 (12.34%) | 208.00 (12.44%) | 74.00 (12.07%) |  |
| Sepsis3, N (%) |  |  |  |  |
| No | 1,601.00 (70.07%) | 1,083.00 (64.77%) | 518.00 (84.50%) | <0.001 |
| Yes | 684.00 (29.93%) | 589.00 (35.23%) | 95.00 (15.50%) |  |
| CRRT, N (%) |  |  |  |  |
| No | 1,949.00 (85.30%) | 1,534.00 (91.75%) | 415.00 (67.70%) | <0.001 |
| Yes | 336.00 (14.70%) | 138.00 (8.25%) | 198.00 (32.30%) |  |
| AKI, N (%) |  |  |  |  |
| No | 376.00 (16.46%) | 322.00 (19.26%) | 54.00 (8.81%) | <0.001 |
| Yes | 1,909.00 (83.54%) | 1,350.00 (80.74%) | 559.00 (91.19%) |  |
| Ventilation, N (%) |  |  |  |  |
| No | 530.00 (23.19%) | 421.00 (25.18%) | 109.00 (17.78%) | <0.001 |
| Yes | 1,755.00 (76.81%) | 1,251.00 (74.82%) | 504.00 (82.22%) |  |
| ECMO, N (%) |  |  |  |  |
| No | 2,269.00 (99.30%) | 1,667.00 (99.70%) | 602.00 (98.21%) | <0.001 |
| Yes | 16.00 (0.70%) | 5.00 (0.30%) | 11.00 (1.79%) |  |
| Vp, N (%) |  |  |  |  |
| No | 1,037.00 (45.38%) | 839.00 (50.18%) | 198.00 (32.30%) | <0.001 |
| Yes | 1,248.00 (54.62%) | 833.00 (49.82%) | 415.00 (67.70%) |  |

Abbreviation: NMBP Non-invasive mean blood pressure ; NDBP Non-invasive diastolic blood pressure ; Rdw red cell distribution width; INR International normalized ratio; Pt Prothrombin time; APACHEII Acute Physiology and Chronic Health Evaluation II; IABP Intra-aortic balloon pump; CRRT Continuous renal replacement therapy; AKI Acute kidney injury; ECMO Extracorporeal Membrane Oxygenation; Vp vasopressin.

### Characteristics of AMI in the Survival and Death Groups

In ICU-admitted patients with AMI, there were statistically significant differences between the survival and mortality groups in terms of age, initial vital signs (including systolic blood pressure, diastolic blood pressure, heart rate, pulse oxygen saturation), and initial laboratory test indicators (such as hematocrit, platelet count, hemoglobin, red cell distribution width, etc., totaling 36 variables). In addition, there were statistical differences between the two groups in terms of disease severity scores (SOFA score, APACHE II score), the incidence of comorbidities (sepsis, acute kidney injury), and the proportion of patients receiving specific therapeutic interventions (intra-aortic balloon counterpulsation, extracorporeal membrane oxygenation, continuous renal replacement therapy, mechanical ventilation, and the use of vasoactive drugs).Table 1 shows the above results in detail.

### Screening Predictors of Mortality Risk in ICU Patients with AMI

Fifteen potential predictors of 28-day mortality in AMI were selected by Least Absolute Shrinkage and Selection Operator (LASSO) regression, including age, heart rate, WBC counts, absolute lymphocyte counts, lactate, creatine kinase isoenzymes, total calcium, blood sodium, alanine aminotransferase, urea nitrogen, fluid balance, SOFA score, APACHEII score, whether hemopurification therapy and whether using vasoactive drugs (Figs. 2 and 3). Supplementary Table 1 presents the Lasso binomial regression Lambda value coefficient table.

**Figure 2.**
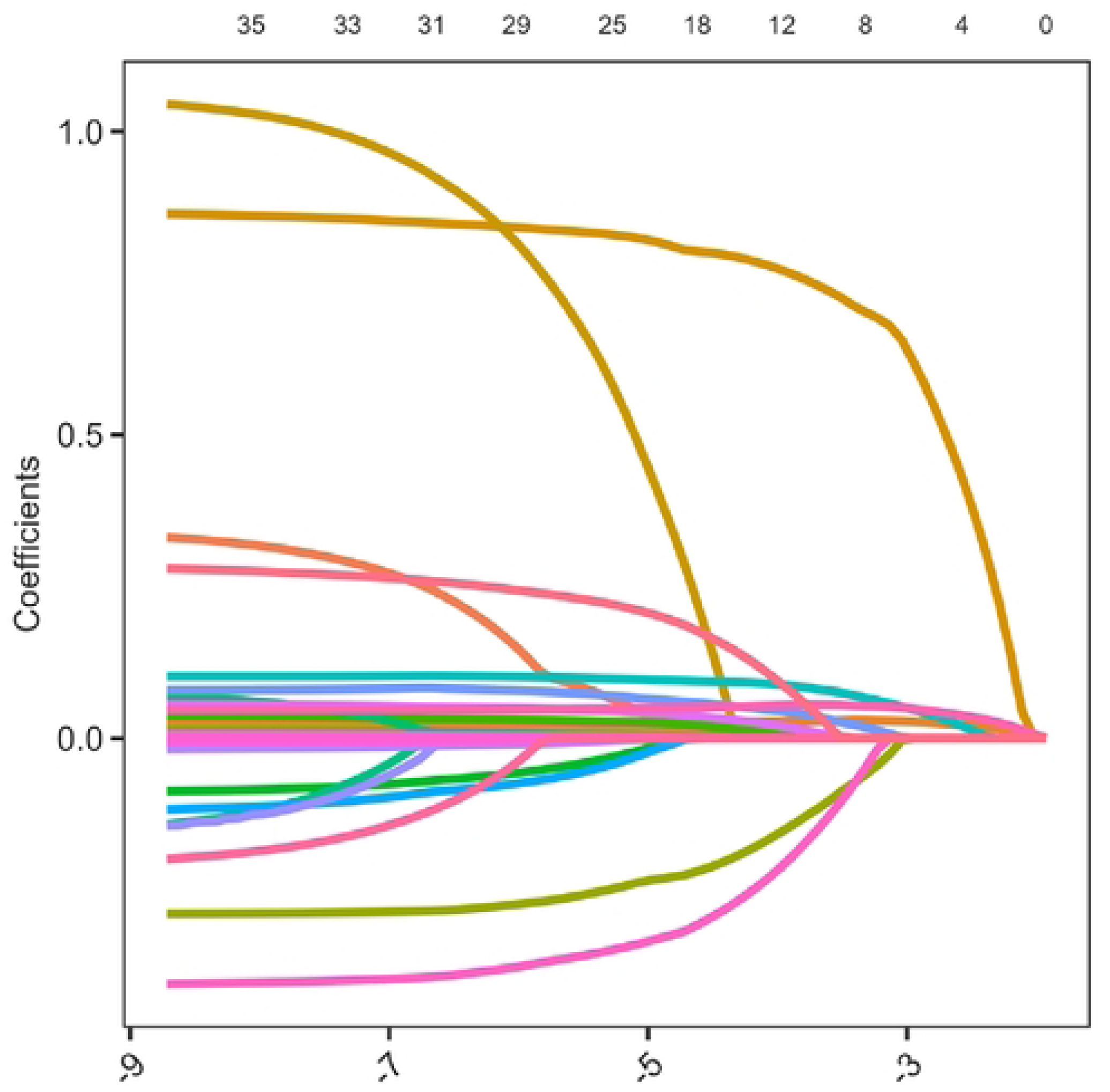
Lasso regression trajectories for each variable.

**Figure 3.**
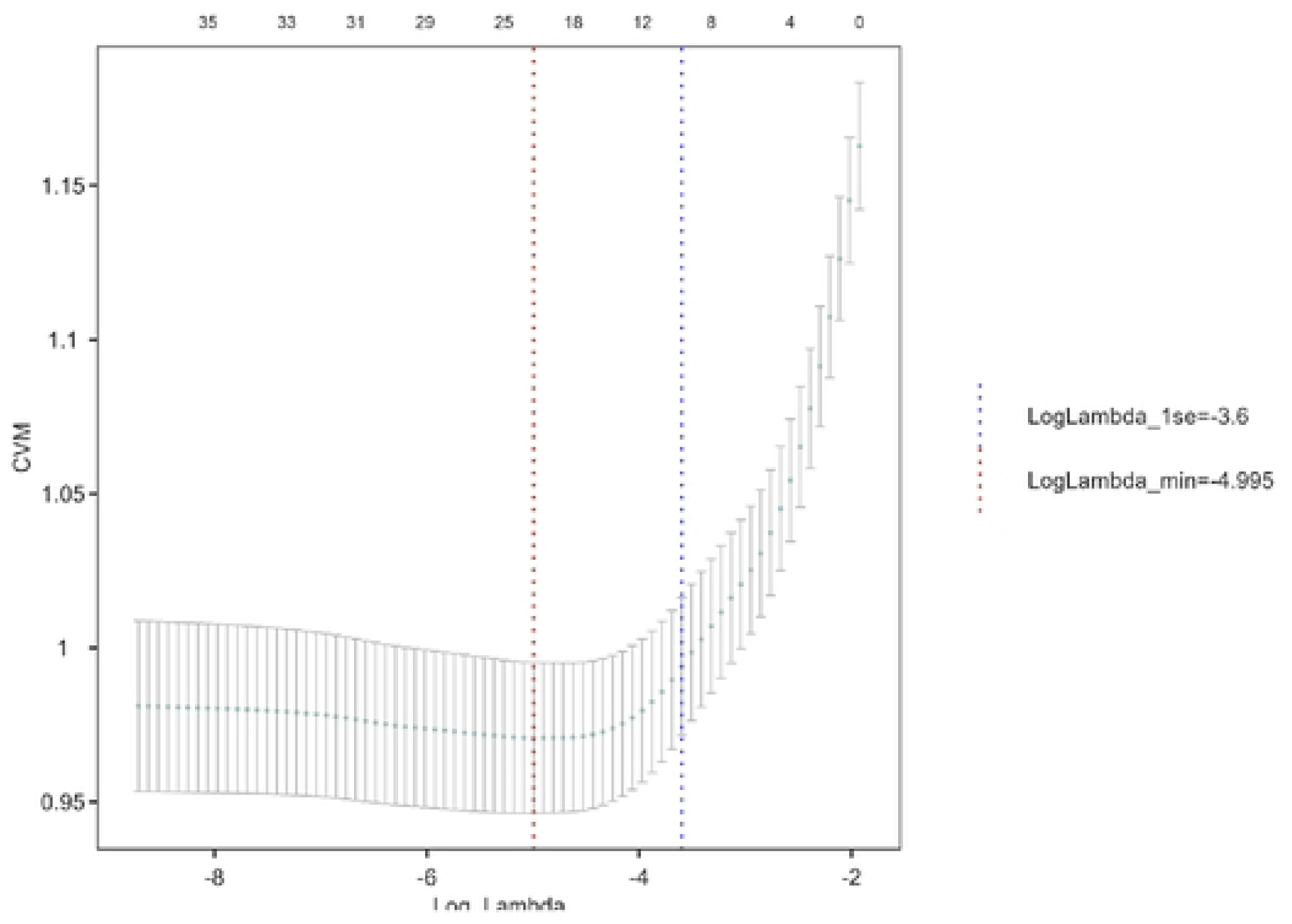
Cross-validation (CV) plot for Lasso Regression.

### Model Development and Performance Evaluation

Ten prediction models were constructed using machine learning algorithms, including Adaptive Boosting(AdaBoost TEST), Categorical Boosting(GatBoost TEST), Decision Tree(Desicion Tree TEST), Gradient Boosting(GBDT TEST), K-Nearest Neighbors(KNNC TEST), Light Gradient Boosting(LGBM TEST), Logistic Regression(Logistic TEST), Random Forest(RFTEST), Support Vector Machine(SVMTEST), and Extreme Gradient Boosting(XGBTEST). In the machine model, according to the area under the AUC curve, the three prediction models show good predictive performance for the risk of ICU death from AMI(Fig.4). The CatBoostTEST model has an AUC=0.78, the LGBMTEST model has an AUC=0.766, and the RFTEST model has an AUC=0.755.

**Figure 4.**
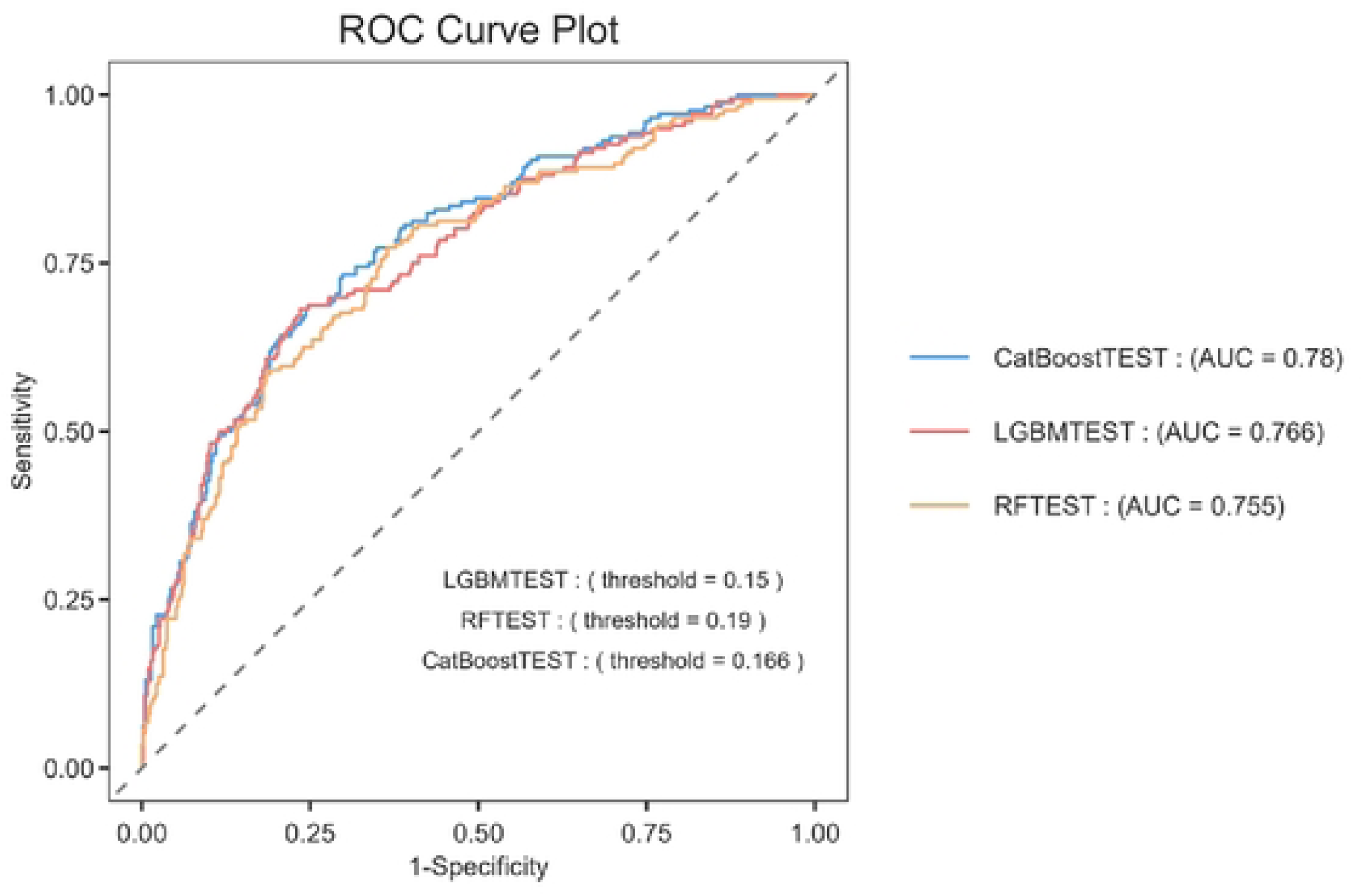
Receiver Operating Characteristic (ROC) curves for the Categorical Boosting (GatBoost TEST), Light Gradient Boosting (LGBM TEST), and Random Forest (RFTEST) are presented to predict the probability of ICU mortality risk in patients with AMI.

### SHAP analysis on extreme gradient boosting

To further calculate the contribution value of each feature in the prediction model, and to understand which features are the most critical and whether they have a positive or negative impact on the prediction results, we applied SHAP(SHapley Additive exPlanations) analysis. Fluid balance, lactate dehydrogenase, red cell distribution width had the greatest influence on model output. SOFA score, lactate, troponin T, albumin, age, glucose, APACHEII score had a consistent influence on model output (Fig.5). Elevated lactate, glucose, lactate dehydrogenase are associated with an increased risk of death in ICU AMI patients. Negative fluid balance is associated with a decreased risk of death in ICU AMI patients (Fig.6).

**Figure 5.**
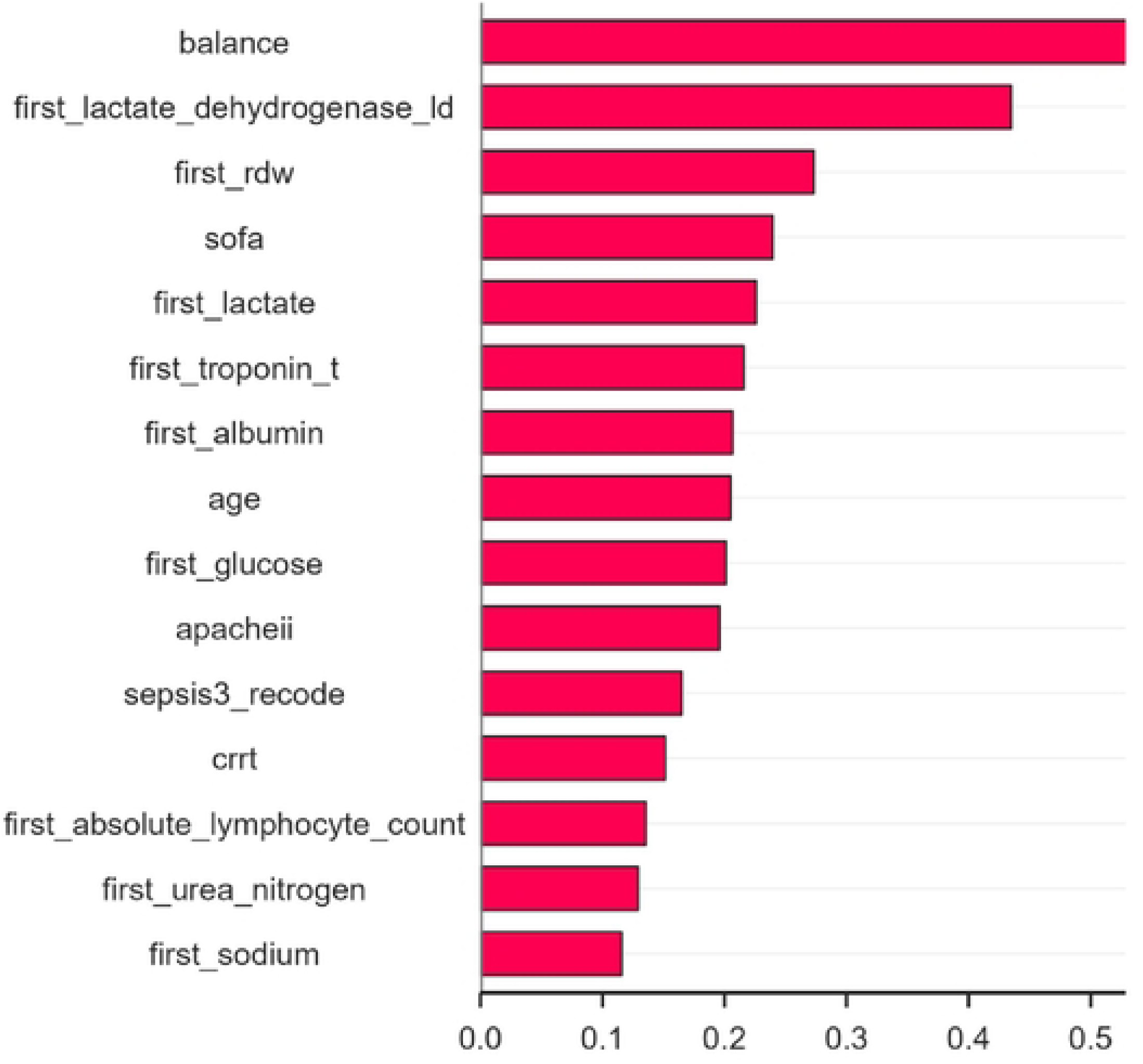
Predictive feature importance ranking for critical outcomes

**Figure 6.**
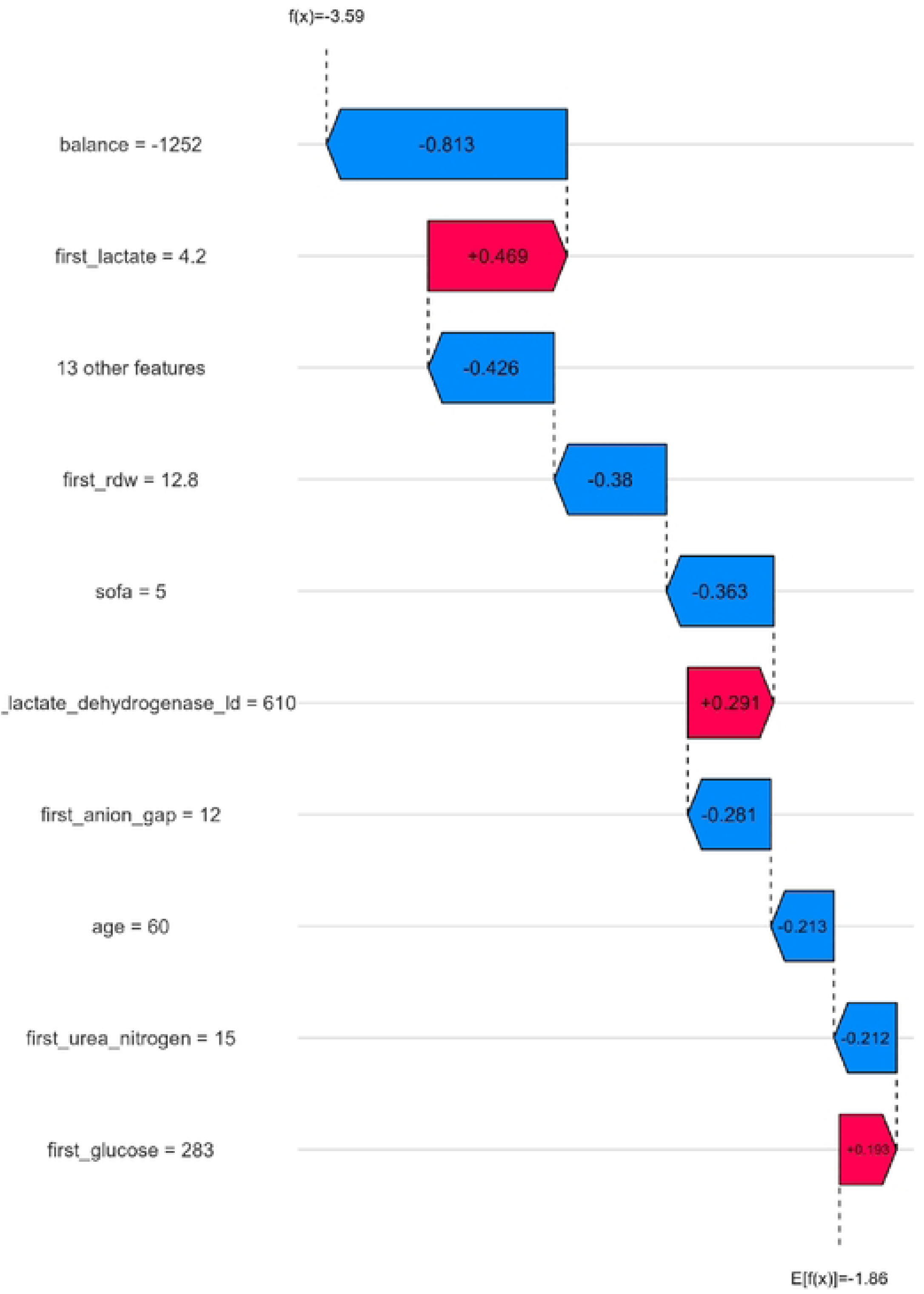
The SH.AP analysis shows the value and direction of the contribution of each feature to the prediction.

## Discussion

AMI as one of the leading causes of cardiovascular disease mortality worldwide, has always posed a significant challenge in clinical practice due to its high mortality rate in ICU[18]. Based on the data of 2285 AMI patients from the MIMIC-IV database, this study constructs an ICU mortality risk prediction model through machine learning algorithms, in conjunction with SHAP analysis, uncovers key influencing factors, providing new theoretical foundations and practical tools for clinical decision-making. Patients with AMI in the ICU have complex and variable conditions, often accompanied by complications such as multiple organ dysfunction and infections[19–20]. Traditional scoring methods struggle to dynamically assess risk in real-time. The results of this study indicate that the machine learning model constructed based on 15 selected variables has good predictive performance for the mortality rate of AMI patients in the ICU. Among them, the AUC of the CatBoostTEST model reached 0.78, while the AUCs of the LGBMTEST and RFTEST models were 0.766 and 0.755 respectively, significantly outperforming traditional risk scoring systems. This discovery further validates the superiority of machine learning technology in predicting the prognosis of critical cardiovascular conditions, offering a new perspective to address the limitations of traditional scoring systems.

The SHAP analysis, as an important tool in explainable machine learning, quantifies the contribution and direction of influence of each variable on model output, providing intuitive evidence for understanding the core mechanisms of ICU mortality risk in AMI patients[21–22]. Our results show that fluid balance, lactate dehydrogenase, and red cell distribution width are the primary factors affecting model output, while SOFA score, lactate, troponin T, age, blood glucose, APACHE II score also have stable contributions. These findings are highly consistent with pathophysiological mechanisms and clinical practices. Fluid balance has the highest weight in the SHAP analysis, and negative fluid balance is associated with a reduced mortality risk. This result highlights the central position of fluid management in the ICU. For AMI patients, especially those with combined heart failure or cardiogenic shock, fluid overload can lead to pulmonary edema, increased cardiac preload, and increased myocardial oxygen consumption, further exacerbating cardiac dysfunction; whereas moderate negative fluid balance (while maintaining circulatory stability) can reduce cardiac burden and improve oxygenation and organ perfusion[23–24]. In clinical practice, fluid management in AMI patients needs to strike a precise balance between maintaining effective circulation and avoiding volume overload[24]. The SHAP analysis results provide data support for this balance, suggesting that fluid balance monitoring should be a routine focus in ICU management for AMI patients. Elevated lactate dehydrogenase is associated with an increased mortality risk, which may be related to myocardial cell necrosis, hemolysis, or liver injury[25]. Lactate dehydrogenase is widely present in tissues such as the myocardium, liver, and kidneys. During an AMI, the release of lactate dehydrogenase from necrotic myocardial cells and factors like hypoxia and inflammation can further damage other organs, keeping its levels elevated[26]. The high weight of red cell distribution width reflects the body’s chronic inflammation and nutritional status. Its increase might be associated with prolonged hypoxia, abnormal iron metabolism, or suppressed bone marrow hematopoietic function, all of which can exacerbate the pathophysiological disturbances in AMI patients and increase mortality risk[27]. Elevated lactate is a direct marker of tissue hypoxia and metabolic. In AMI patients, its levels are significantly correlated with infarct size, cardiac function grading, and the incidence of shock; elevated blood glucose might be related to stress hyperglycemia[28]. High blood glucose can exacerbate myocardial injury and poor prognosis through mechanisms such as oxidative stress induction, endothelial dysfunction, and increased infection risk[30]. This suggests that close monitoring and control of lactate and blood glucose levels might become important targets for improving prognosis in clinical management.

The stable contributions of SOFA score and APACHE II score in the SHAP analysis once again emphasize the importance of multi-organ function protection. The death of AMI patients is often not caused by a single myocardial injury, but the terminal result of multi-organ failure. For example, myocardial infarction can lead to acute kidney injury through cardiogenic shock, causing insufficient renal perfusion, and respiratory failure through hypoxia and inflammatory response[31–33]. These chain reactions are all captured by SOFA and APACHE II scores, which in turn affect the model’s judgment of mortality risk. The impact of age and troponin T further verifies the clinical rationality of the model[34–35]. The decline in physiological reserve associated with aging makes elderly patients more vulnerable to the impact of AMI[36]. Troponin T, as a highly sensitive marker of myocardial injury, directly reflects the degree of myocardial necrosis and is one of the gold standards for prognostic assessment[37]. Their high weight in the SHAP analysis ensures the consistency of the model with traditional clinical understanding and enhances the credibility of the results. The results of this study are highly consistent with the research trends of machine learning in the field of cardiovascular critical illness in recent years, and have unique innovations in terms of data integration and model interpretability.

However, this study also has limitations. Firstly, the research is a retrospective design, and inevitably there is a selection bias. The patients in the MIMIC-IV database mainly come from European and American countries, which may differ from the clinical characteristics of the Chinese population, and the extrapolation of the model needs to be verified in domestic multicenter data. Secondly, the model did not include dynamic change data. The physiological indicators of ICU patients (such as lactate, blood pressure) change dynamically over time, while this study only uses baseline data at admission, which may omit important prognostic information. Finally, the model lacks external validation. This study only developed models based on the MIMIC-IV database, and has not been validated in an independent cohort, and its stability and universality need further confirmation. Despite certain limitations, this study still provides an important reference for the refined management of AMI patients. In the future, through multicenter validation, dynamic data integration, and interventional research, there is hope that machine learning models will be more widely applied in clinical practice, ultimately achieving the goals of reducing the ICU mortality rate of AMI patients and improving their prognosis.

## Conclusion

The prediction model of ICU mortality rate for AMI patients constructed by machine learning algorithms in this study demonstrates good predictive performance.

## Data Availability

MIMIC database: https://physionet.org/content/mimiciv/3.1/.

## Acknowledgments

Not applicable.

## Authors’ contributions

All authors contributed substantially to the study design, data interpretation, and the writing of the manuscript. Ying-mei Xiao contributed to the study design. Ying-mei Xiao, Juan Zhang, Mao-juan Wang contributed to Data Analysis and Manuscript Writing. All authors reviewed the manuscript. The author(s) read and approved the final manuscript.

## Availability of data and materials

MIMIC database: https://physionet.org/content/mimiciv/3.1/.

## Consent for publication

This study utilized the MIMIC-IV database, a publicly available, de-identified database. As this study involved the analysis of pre-existing, anonymized data, separate ethics approval for this specific analysis was not required.

---

**Supplementary table 1.**
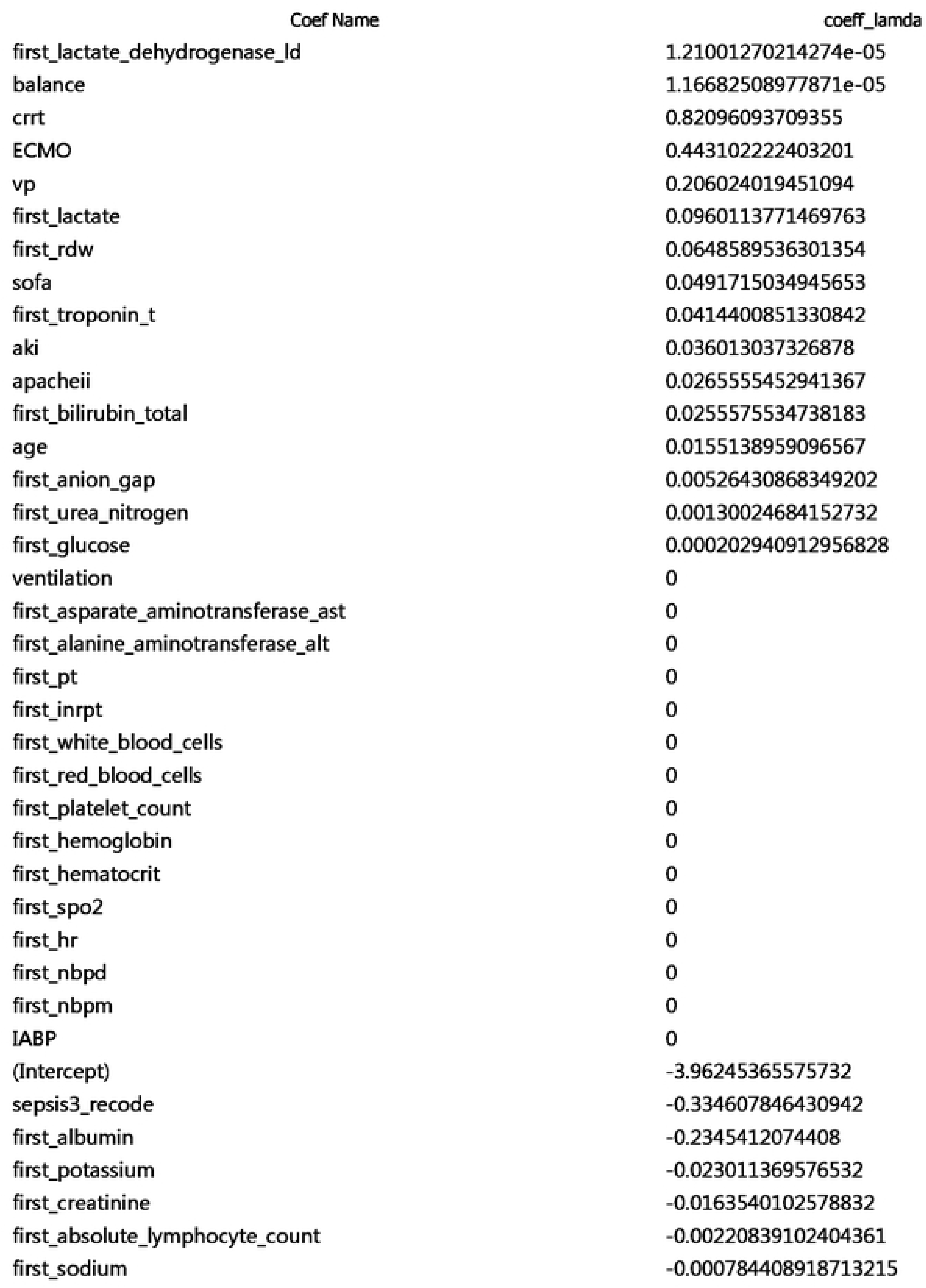

